# School staff and university instructors’ perceptions of barriers and facilitators of whole-grain consumption among children in Thailand: qualitative findings

**DOI:** 10.64898/2026.09.22.26363746

**Authors:** Nutnicha Paisanvit, Punnee Ponprachanuvut, Kantanit Chammari, Nachon Raethong, Nuttarat Srisangwan, Arisa Keeratichamroen, Shuqi Wang, Hoang Tong, Rajesh Kumar Rai, Warangkana Srichamnong, Sabri Bromage, Mika Matsuzaki

## Abstract

Thailand is experiencing a rapid rise in childhood obesity, increasing the risk of early-onset non-communicable diseases. Although whole grains support weight management and reduce non-communicable diseases risk, consumption remains low in Thailand and is insufficiently addressed in the Thai Food-Based Dietary Guidelines. Whole grains are also rarely included in the National School Lunch Program. Given its high participation rate, the school lunch program represents a strategic platform for increasing whole grain intake and fostering lifelong healthy eating habits. This study aimed to identify facilitators and barriers to increasing whole grain provision in Thai elementary schools from the perspectives of adult stakeholders

We conducted in-depth interviews guided by the Integrated Behavioral Model with 19 teachers and contracted school cooks from two Thai elementary schools, and with 10 faculty and staff members at Mahidol University. Interviews explored perceived norms regarding student whole grain consumption, attitudes toward whole grain, and perceived facilitators and barriers to whole grain provision in schools. Data were analyzed using framework analysis, combining the Integrated Behavioral Model-informed thematic categorization with inductive coding. Transcripts were independently coded by two researchers to develop an Integrated Behavioral Model-grounded conceptual framework describing factors influencing intentions to increase whole grain provision in Thai elementary schools.

Participants were predominantly female (93.1%) and highly educated (89.7% with a bachelor’s degree or higher). Most supported incorporating whole grains into school meals, citing their health benefits for schoolchildren. However, participants reported low WG intake among schoolchildren, attributable to limited availability at both schools and homes. Two primary barriers to whole grain provision were identified. The first was student-level resistance, including unfamiliarity and sensory aversions, particularly toward brown rice. The second involved systemic challenges, including the absence of regulatory guidance promoting whole grain inclusion and operational constraints related to contracted food services and inflexible menu systems. Multi-level engagement among schools, households, and government agencies was identified as a key facilitator of increased whole grain provision in school meals. These findings indicate that although teachers and cooks are receptive to whole-grain provision in school meals, translating this support into practice will require coordinated, multisectoral action to address persistent structural barriers at the student, school, and policy levels. Future research should incorporate children’s perspectives and examine a broader range of school settings to inform the design of effective, sustainable strategies for whole grain integration.

## Introduction

Childhood overweight and obesity are significant public health issues in Thailand. Between 1995 and 2014, overweight/obesity prevalence more than doubled among Thai school-aged children (6-14 years), from 5.8% to 13.9% [1]. Childhood obesity is expected to persist and intensify globally, with Southeast Asia projected to have one of the largest increases in childhood obesity prevalence by 2050 [2]. The rise of childhood overweight/obesity is concerning, as overweight/obesity is likely to persist into adulthood, potentially causing both immediate health consequences at a young age and increased risk of earlier onset of non-communicable diseases (NCDs) such as type 2 diabetes mellitus and cardiovascular diseases, later in life [3] [4].

Whole grains (WG) are rich in dietary fiber, vitamins, minerals, and phytochemicals, which contribute significantly to overall health and well-being. Studies have consistently shown a strong correlation between regular WG consumption and a reduced risk of weight gain and NCDs [5–7]. In Thailand, whole grains are mostly found in pigmented or minimally milled rice varieties, including brown rice, Riceberry, Sangyod, Leum Pua, Hom Nil, Sinlek, and Red Hom Nil [8]. These varieties reflect decades of agricultural and food science innovation. Despite rice being the primary staple, the national data indicate extremely low consumption of brown rice among children aged 6–12.9 years (5.67 g/day), with no reported intake of other whole-grain sources [9]. Currently, whole grains are not prioritized in Thai regulations, policies, or public health programs. The national Thai Food-Based Dietary Guidelines (Thai FBDGs) simply encourage the consumption of brown rice, without mandating or providing specific recommendations for it, nor mentioning other types of WG [10].

In 2020-2021, over 3 million primary school students received food through school meal programs in Thailand [11]. The national school lunch program, while focusing on providing nutritious meals, currently does not specifically mandate or encourage the inclusion of WG in school meals [10,12]. Given the high attendance rate (94.8% in 2023), this setting offers an ideal venue for schoolchildren to increase WG consumption and develop lifelong healthy eating habits to prevent NCDs [13,14]. In the United States, where WG inclusion is required in the National School Lunch Program and Breakfast Program, studies have shown increases in WG consumption among youth, with the evidence suggesting specifically the beneficial effects of this school nutrition policy [15].

Given the potential health benefits of including WG in National School meals in Thai elementary schools, it is essential to understand the potential facilitators and barriers involved to realize this change. This study was conducted alongside our survey on knowledge and perceptions around whole-grain consumption among children in Thailand at the same elementary schools to provide complementary insights that can contribute to advocacy for WG provision in school meals and further increase WG consumption among Thai children.

However, to the best of our knowledge, no studies have specifically explored these aspects. This study aimed to address a critical knowledge gap by identifying the factors influencing the adoption of WG in school meals from the perspectives of adult stakeholders. Specifically, we aim to 1) explore perceptions towards introducing WG in school meals among the school staff and university-level nutrition instructors, and 2) identify *barriers* and *facilitators* to increasing WG consumption in elementary schools in Thailand perceived by the school staff and university Instructors. Both aims were achieved through qualitative analysis of stakeholder interviews.

## Methodology

This study employed a qualitative descriptive approach, grounded in a constructivist/interpretivist paradigm, which assumes that knowledge and meaning are co-constructed through participants’ lived experiences and perspectives [16]. This paradigm was selected because the study sought to explore multiple, context-specific realities regarding whole-grain integration in Thai elementary schools, where stakeholder perspectives and social context are central to understanding barriers and facilitators.

### Participants

Participants in this study comprised two groups recruited from elementary schools and a university in Nakhon Pathom, Thailand. The first group included teachers from grades 4–6 and hired cooks under contract from two participating elementary schools (School 1 and School 2). These schools were purposively selected because they participated in a government-subsidized lunch program with mandated nutritional standards and had collaborated with the Institute of Nutrition, Mahidol University (INMU). At the time of the study, neither school had an existing health education program. The second group included university personnel, comprising faculty and academic staff from INMU. These participants were included to provide expert perspectives on the Thai food system and insights into the practical integration of whole grains (WG) into school-aged children’s diets. The eligibility criteria for this study were:

1) 18 years of age or older at the time of the study
2) proficient in either Thai or English
3) willingness to participate and were able to complete the IDI
4) consented to have their interview recorded and transcribed

### Recruitment procedure

Purposive sampling was used to recruit INMU staff from November 6, 2023 – February 13, 2024 and school staff on February 15, 2024. Fifteen teachers and cooks from School 1 and fourteen from School 2 were approached in person at their respective schools. The schoolteachers and cooks were purposively recruited based on their direct roles in the school food environment, including designing school menus and/or preparing and cooking school lunches for students. A total of 144 university faculty and staff were invited via email. The university personnel were purposively recruited based on their expertise in nutrition and the food system. No incentives were provided for participation. In total, 19 teachers and cooks from the two schools and 10 university faculty and staff were enrolled. Although the sample size was determined a priori, data saturation was assessed iteratively throughout the coding process. Little or no additional information emerged after the 12th interview, suggesting that saturation had been achieved. This is consistent with prior qualitative research demonstrating that data saturation in individual in-depth interviews is commonly reached within the first 10 interviews [17,18]. Details on sample recruitment, representativeness, and handling of missing data are provided in Additional File 1.

### Data Collection

The In-depth interview (IDI) guide was designed based on the Integrated Behavioral Model (IBM) [19]. The IDI guides for subjects primarily focused on attitudes, perceived norms, and personal agency, including perceived control and self-efficacy. Additionally, participants were asked to identify barriers and facilitators to the incorporation of WG in Thai school settings. For INMU faculty/staff, questions on their attitudes toward the health benefits of WG, the sensory properties of WG, and their influence on schoolchildren’s perception and/or behavior were excluded. Instead, we focused on their attitude toward WG consumption among Thai children and their perspective on increasing WG consumption in Thai elementary schools. We also specifically asked personal agency-related questions, including their ability to identify WG and WG products and their skills in preparing and cooking WG, to schoolteachers and cooks.

Interviews with school staff were conducted in person during school hours. Interviews with the university faculty and staff were conducted either in person or virtually, depending on participant availability. All interviews were conducted between January 2024 and February 2024 by trained INMU researchers. Interviews were audio-recorded using trained researchers’ smartphones, transcribed by the bilingual researcher on the study team, and translated from Thai into English using Microsoft Word’s built-in AI translation tool [20], followed by manual refinement and verification by native speakers to maintain the integrity and nuances of the original participant narratives. Written consent forms were obtained before data collection. The study protocol was approved by the Mahidol University Central Institutional Review Board (COA No. MU-CIRB 2023/186.2212) and the Johns Hopkins Bloomberg School of Public Health Institute of Review Board (IRB No:26258).

### Data Analysis

IDI data were analyzed following a framework analysis approach with theme categorization informed by the IBM [19]. The primary coder (NP) first became familiar with the data by thoroughly reviewing both the original Thai transcripts and their English translations [19]. The primary coder then conducted inductive coding across three of the richest transcripts to identify emerging themes not anticipated by the IBM framework [19]. The emergent themes were categorized into subthemes under each element of IBM, forming a structured codebook [19]. Codes that did not align with the IBM framework were placed under an “Others” category.

To ensure comprehensiveness and clarity, the codebook was reviewed and refined twice: first by two independent reviewers (SW and HT), and then by the main coders (NP and SW). After the first codebook validation, 30 codes were kept under 7 main themes. For the second validation, the codebook was piloted independently by the two coders on six transcripts before being applied to the remaining data. Following codebook validations, the remaining transcripts were divided equally between NP and SW for indexing and analysis using Atlas.ti software [21]. Codes were applied through contextual interpretation, with perceptions categorized as facilitators or barriers based on their role in the discussion rather than predefined keywords. After coding, the team reorganized codes and themes to reduce redundancy and confirm within-theme coherence. The final, IBM-grounded conceptual framework was established, depicting factors that influence intentions to improve WG provisions in Thai elementary schools, as appears in Figure 1. To enhance trustworthiness, an audit trail of analytical decisions was maintained throughout the coding process. Member checking was not conducted due to logistical constraints; however, credibility was strengthened through independent coding, iterative codebook refinement, and ongoing reflexive discussion among the research team [22]. The SRQR checklist is provided in Additional File 2.

**Fig 1.**
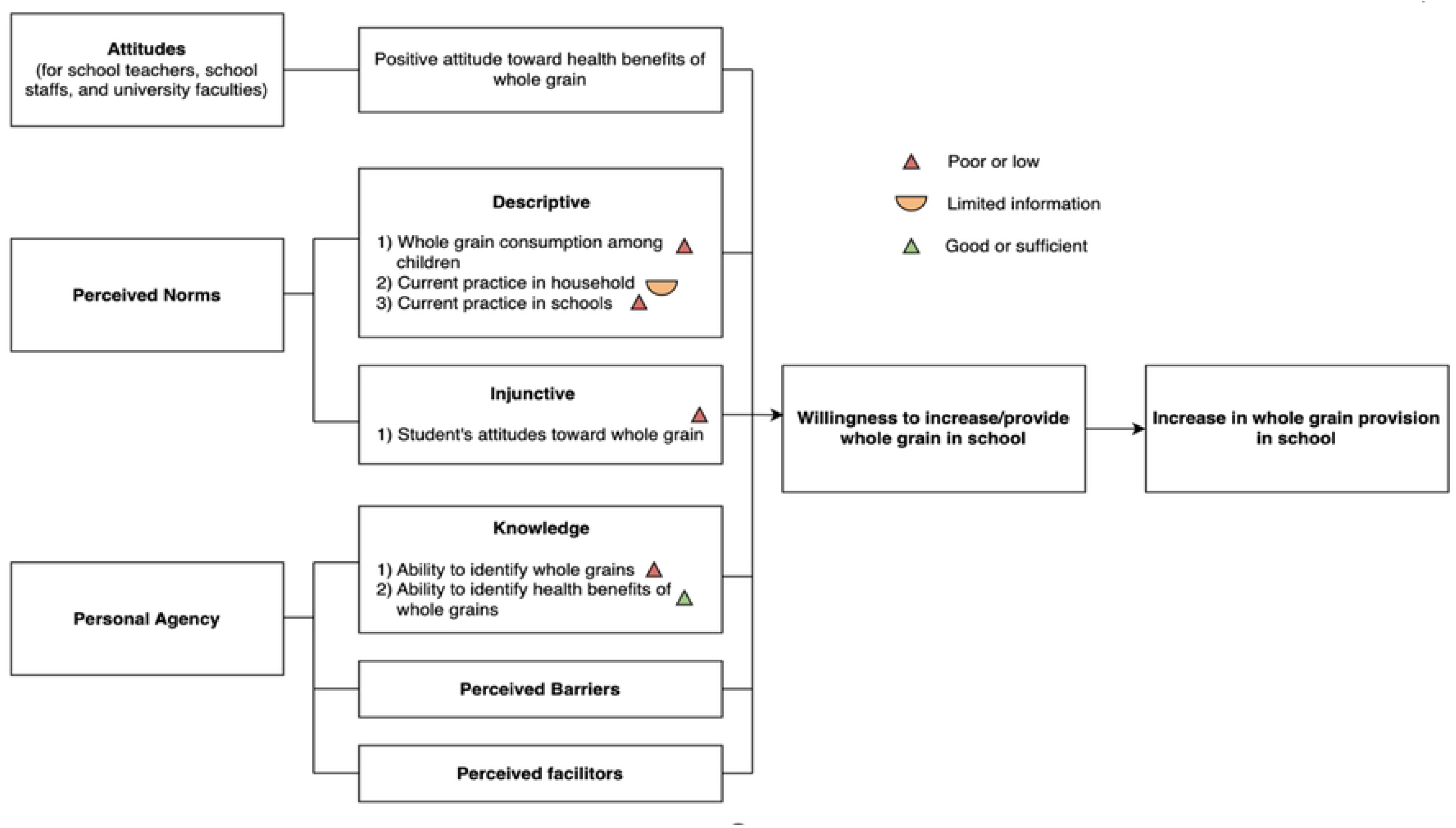
Conceptual Framework: Factors Influencing Improvement of WG Provisions in Thai Elementary Schools.

### Reflexivity

The majority of the research team consisted of Asian females with academic training spanning from bachelor’s to doctoral degrees in nutrition and public health. Most team members grew up in Thailand or other Asian countries where rice serves as a primary dietary staple, providing shared cultural familiarity with rice-centric food environments and the limited presence of whole grains in habitual diets. This cultural proximity may have facilitated contextual understanding of participant perspectives but may also have introduced assumptions about dietary norms that were actively monitored throughout analysis. Half of the team members brought institutional knowledge with direct expertise in the Thai school lunch system (PP, KC, NR, NS, AK, WS). RKR, SB, and MM offered additional experience in survey design and methodology, school lunch interventions, and other public health programs focusing on child nutrition and obesity in Thailand and international settings.

All interviews were conducted by PP, KC, NR, NS, and AK (with oversight on question content from SW). All interviewers were native Thai speakers with expert familiarity with the Thai school lunch context, facilitating linguistic authenticity and cultural rapport. Translation was conducted by NP, a bilingual Thai-English researcher and native Thai speaker, who manually verified and refined the translations to preserve the cultural nuances of participant narratives. We acknowledge that institutional affiliation with Mahidol University and Johns Hopkins may have introduced social desirability bias in participant responses, and that the team’s nutrition training may have shaped interpretive assumptions regarding whole-grain benefits. To mitigate this, all researchers documented assumptions, reflected their positionality, and set aside prior views to the best of their ability throughout data collection, coding, and interpretation.

## Results

We interviewed 29 participants (Table 1). The participants were primarily female (93.1%) and had bachelor’s degrees or higher (89.7%).

**Table 1:** Sociodemographic characteristics of the interview participants at elementary schools and at a university.

|  | **School 1** | **School 2** | **University** |
| --- | --- | --- | --- |
| **Number of participants (n, %)** | 4, 14% | 15, 52% | 10, 34% |
| **Age (years), mean±standard deviation** | 49.5±8.9 | 37.1±11.7 | 39.1±12.3 |
| **Age range, (years)** | 38-57 | 18-54 | 24-55 |
| **Sex (n)** |  |  |  |
| **Female** | 4 | 14 | 9 |
| **Male** | 0 | 1 | 1 |
| **Education level (n)** |  |  |  |
| **High school**<br>**certificate**<br>**equivalent or less** | 0 | 3 | 0 |
| **Bachelor's degrees** | 1 | 9 | 4 |
| **Master's degrees** | 3 | 3 | 6 |

Five themes were identified and organized into three overarching concepts: stakeholder perceptions of WG provision in schools (Theme 1); structural and behavioral barriers shaping current WG consumption (Themes 2–4); and system-level facilitators to support sustainable WG integration in school meals (Theme 5).

### Theme 1. Favorable perception toward increasing WG in schools

Most participants were favorable toward including WG in school meals, frequently citing them as a healthier alternative with superior nutritional benefits and emphasizing their potential to improve students’ dietary quality.

> *“If it is unpolished rice, it will be more beneficial than polished rice, that is, it will have vitamins and proteins, and the child will get more complete nutrients.” (School staff 1)*

Nearly half of educators also viewed WG promotion as consistent with their broader goals of fostering healthier eating habits among students and expressed openness to implementation. While there were concerns about potential implementation challenges, some participants believed that students would adjust over time without significant resistance.

### Theme 2. Low WG consumption and availability among Thai elementary schoolchildren at school and at home

About half of participants stated that students rarely consume WG. The availability of WG products remained limited in school settings, both in school lunch programs and in school shops. Brown rice was noted as largely absent from school meals, while white rice was the staple. WG snacks were also rarely offered, with only occasional sales of whole-wheat bread at School 2.

> *“At schools, they barely, almost never, eat brown rice because we provide them with white rice. When the children eat bread, they eat white bread and not whole wheat.” (School staff 2)*

Some participants noted that children from health-conscious families were more likely to consume brown rice, often mixed with white rice, when it was served at home. However, limited exposure to WG at home was also reported. The participants noted that students raised primarily on white rice lacked exposure to WG, leading to low awareness of grain processing and WG-related nutritional benefits, which may further contribute to hesitancy or rejection.

> *“If we ask them whether they eat [unpolished rice], they say no. When asked why, they say their home doesn’t cook it, their parents don’t eat it.” (University staff 1)*

Although neither school offered brown rice in regular meals, both schools occasionally offered WG in alternate forms, such as corn and Job’s tears in cold desserts. Introducing mixed brown and white rice at school was welcomed by students, as noted by a School 1 teacher. However, a School 2 teacher reported lower intake when WG was served.

### Theme 3. Dislikes and unfamiliarity with WG as a barrier to increasing WG consumption among schoolchildren

Participants identified schoolchildren’s resistance, unfamiliarity, and sensory aversions as major challenges to increasing WG consumption in schools. Generally, many participants felt that students found WG products unappealing, particularly brown rice and whole wheat bread. Many noted WG characteristics of coarse texture, dry and greasy mouthfeel, bland flavor, and darker color are key reasons, as they contrast with students’ preferences for soft, sweet, and easy-to-eat foods. Therefore, students often preferred refined grain products over WG when given the option.

> *“From the children’s perspective, they don’t like [WG]. They said that it was crunchy, it was hard, it had a greasy texture, it wasn’t delicious.” (School staff 1)*

However, a few attributed the disinterest to unfamiliarity rather than a strong aversion, given students limited experience with WG. Some participants observed that students were more accepting of whole wheat bread than of brown rice, possibly because whole wheat bread resembles white bread in appearance and texture. A few school staff members reported no observable difference in students’ reactions to WG versus refined-grain products, suggesting that some children may not distinguish between the two. One teacher shared a positive example of students participating in a cooking activity with whole-wheat bread and showing enthusiasm to learn more about WG.

Given the less appealing texture, taste, and appearance of WG, which the children found unfavorable, some participants expressed concern that introducing WG might further reduce their overall food intake. Several educators noted past attempts to introduce brown rice that resulted in substantial food waste.

> *“If it’s rice the kids won’t eat, the schools won’t pass [the performance evaluation] because the children will throw it away.” (University staff 1)*

### Theme 4. Lack of incentives and operational support as barriers to increasing WG in school meals

At the school level, a key barrier to increasing WG consumption among Thai elementary school students was limited availability within the school, which hindered students’ familiarity with WG products. This stemmed from limited educator knowledge of WG benefits.

> *“…right now [educators] still don’t understand why they need to use it or bring WG into the school.” (University staff 2)*

The absence of regulatory guidelines or incentives to promote the serving of WG in schools was frequently noted. Participants noted that schools faced no pressure to proactively provide WG, but efforts were further deterred by perceived student resistance and fear of food waste.

Operational constraints also played a role. As both schools outsourced meal preparation to contractors, any menu adjustments, including WG incorporation, would require negotiation with the contractors, who could object due to budget constraints (22 baht/student/day), higher costs of WG, or ingredient availability. However, one university faculty member suggested that contractors might be more willing to collaborate if they were made aware of the health benefits for students. In addition, a rigid, centralized, district-mandated menu reporting system limited the flexibility to introduce WG recipes, further constraining schools’ ability to adapt menus.

> *“I was only responsible for making the menu…and the contractor will be the one who buys the ingredients. We can determine [the menus], but if the cost is too much, they probably don’t want to change it.” (School staff 3)*

### Theme 5. Multi-level engagement as facilitator for increasing WG in school meals

Participants discussed a wide range of potential facilitators at school and beyond (Table 2). Participants identified households as a critical setting for complementing school-based efforts and improving WG consumption among children, given that students spend the majority of their non-school hours at home. Families were viewed not only as influencers but as facilitators capable of normalizing and maintaining WG consumption. Early home exposure was said to increase openness and reduce resistance at school.

> *“And if the kids don’t eat such grains at home, they won’t eat them here either. Even if they try it here, at home it’s back to the same diet.” (School staff 4)*

**Table 2.**
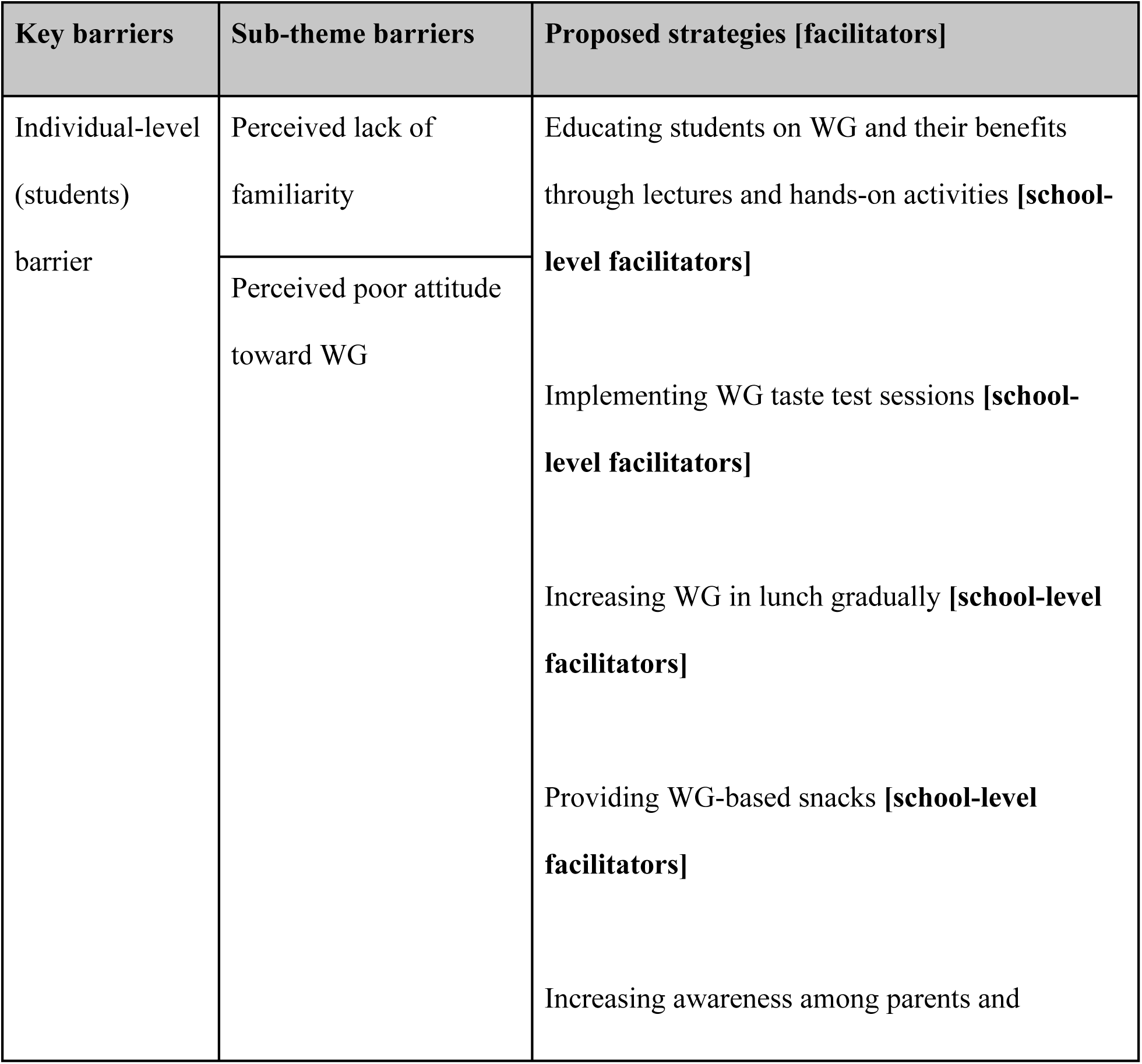
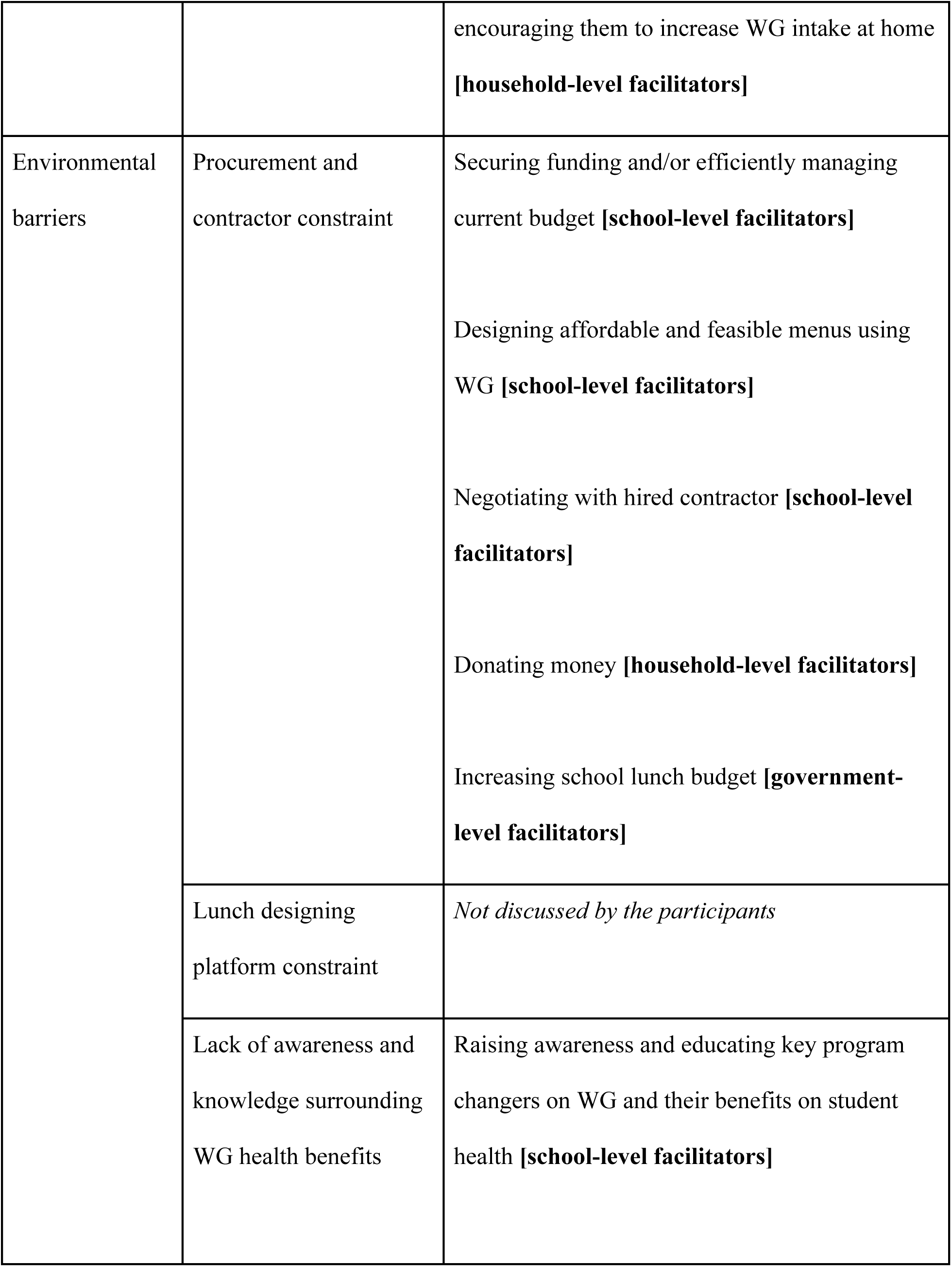
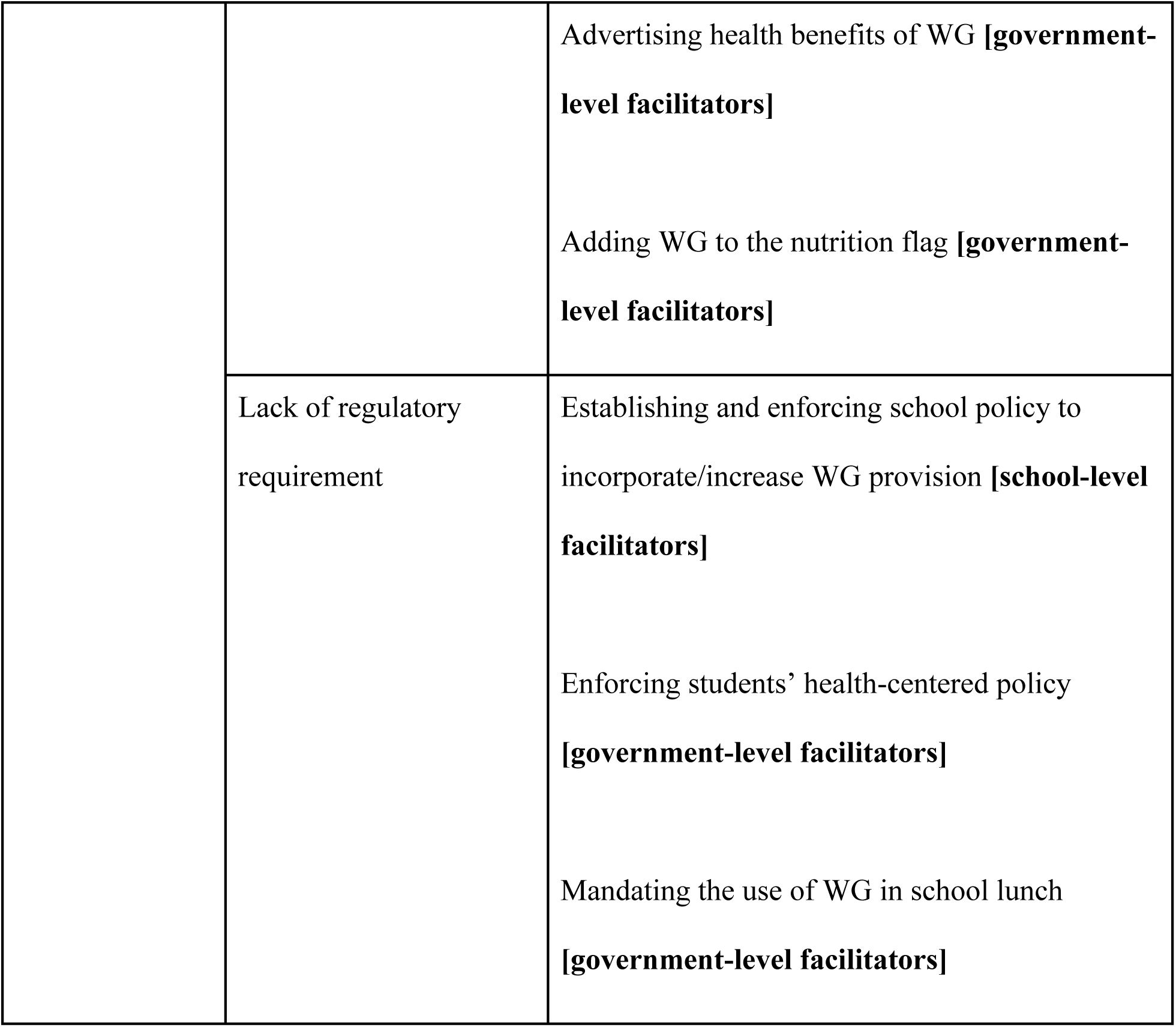
Summary of barriers and their associated proposed strategies and facilitators to increase WG provision in Thai elementary schools.

In cases where schools faced procurement limitations, some participants suggested that household-level efforts could fill the gap by directly introducing WG in packed lunches or making small donations to offset costs. Many participants proposed educating parents about WG, their sources, and their benefits. Increased parents’ awareness of WG could empower them to make informed food choices, model healthy eating behaviors, and reinforce messages children receive at school. While shifting household habits may add expense or inconvenience in the short term, doing so can create a sustainable, health-promoting food environment across settings and support population-level dietary improvement in the long term.

Despite structural and operational barriers, many participants perceived the school as a key platform to increase WG intake by leveraging their control over meal planning, budgeting, and education. Change requires coordinated efforts among health educators, menu planners, and administrators. A university-affiliated faculty member emphasized training these stakeholders on WG and their health benefits to raise awareness and motivate implementation. Several participants also stressed that school- or district-level policies explicitly promoting WG are essential to sustain change by providing ongoing support and clear guidelines.

> *“We need to educate the people who are responsible for the school’s meals [on] what the importance of unpolished grains is…Discuss which menu we can add WG to…From meal planners, budget managers, and suppliers, they also must support each other because some grains can be expensive, so they are not included in the lunch menu.” (School staff 5)*

To mitigate financial constraints, participants suggested that administrative staff could seek additional funding or better allocate existing resources to prioritize WG procurement. Participants also encouraged schools to proactively negotiate with contractors to identify cost-effective and feasible WG options for school availability.

> *“But actually, you can negotiate because it’s for children, it’s done for children. So, if we come up with a menu that has reasonable prices, based on the overall picture.” (School staff 6)*

Schools were also perceived as instrumental in shaping students’ attitudes and behaviors toward WG. Increasing WG exposure and providing education were repeatedly cited to help students familiarize themselves with WG characteristics and potentially reduce their resistance. Some strategies proposed are to gradually increase WG in meals by mixing them with white rice and offering an interactive taste-test session. Regarding education, participants proposed integrating explicit education into the curriculum and reinforcing it through extracurricular activities such as cooking and hands-on projects. Teachers noted that students would be receptive to lessons that were age-appropriate, engaging, and interactive. Several emphasized that elementary grades are a crucial window for promoting long-term dietary behavior change, as students are forming their food preferences and are open to new experiences.

While most participants identified school and household as key facilitators, one participant, who had direct experience working with national nutrition policy, highlighted the potential facilitating role of government-level support, particularly the Ministry of Public Health and the Ministry of Education, as a key facilitator. The Ministry of Public Health, while not directly overseeing school food policy, could mandate WG inclusion through national health campaigns and by updating its nutritional guidance frameworks. In particular, she proposed adding WG to the Thai Nutrition Flag, a widely used guideline for designing balanced school meals. Such inclusion could legitimize WG as a recommended dietary component and drive schools to incorporate it into their lunch programs.

> *“…if we look beyond this, someone with more power than the school director may be the one who sets the budget policy that the school will receive.” (University staff 1)*

The same participant noted that the Ministry of Education, which directly governs school policy and budgeting, could further facilitate change by mandating that a portion of the school lunch budget be allocated to WG procurement. Such a policy could reduce reliance on individual school initiatives and ensure more uniform implementation across the education system.

In addition to these ministerial actions, many proposed broader governmental efforts, such as reducing the price of WG products or improving market availability. These suggestions reflect how government intervention could create an enabling environment for schools to adopt healthier food practices.

For a comprehensive overview, key barriers of WG provision in Thai elementary schools and participants’ proposed strategies for each are further detailed in Table 2.

## Discussion

There was general support for using school meals as a platform to improve knowledge, perceptions, and consumption of WG among Thai schoolchildren. However, participants raised several individual and systemic barriers during the interviews. The primary barriers to introducing WG in school meals included student resistance and unfamiliarity with WG products, and systemic barriers such as resource limitations for school lunch programs and a lack of incentives for schools to offer WG options. To address these barriers, participants suggested a multisectoral approach involving families, schools, and governments in developing a sustainable system to increase WG intake among children through schools in Thailand.

### Individual Barriers

Participants in our study uniformly reported that Thai schoolchildren’s consumption of WG was low. This finding is consistent with reports of low WG intake in other Southeast Asian countries, including Malaysia, Singapore, and the Philippines [23]. Participants mentioned student resistance to switching to WG, with stronger resistance toward rice than other WG products, such as bread. Given that rice is the staple for the majority of the Thai population, this is a key issue to address. Secondly, similar to previous literature, our findings suggest sensory aversions to WG among youth and young adults [24,25]. Studies in Asian populations further indicate that the perceived taste and texture of brown rice pose challenges to increasing consumption [26–29]. Among Southeast Asians, this sensitivity may be linked to habitual white rice consumption, which fosters the ability to detect subtle differences in flavor and texture [30], underscoring the importance of introducing WG early in life to support habitual acceptance.

Participants noted that children’s unfamiliarity with WG is primarily attributed to limited exposure at home and at school, and a lack of knowledge about their health benefits. A study in Nepal found that, despite participants having negative attitudes towards the sensory attributes of brown rice, they became more willing to incorporate it into their diet after trying it in various proportions with white rice [28]. This finding lightened the value of promoting WG provision in Thai elementary schools to help children first become familiar with WG and then be more willing to eat them. Regarding student resistance, one participant expressed concern that serving brown rice might reduce students’ energy intake due to poor acceptability. However, evidence from a school-based WG intervention found no significant difference in energy intake between the intervention and control groups [31]. This suggests that substituting brown rice for white rice may be feasible without compromising total energy consumption.

### Systemic Barriers

Participants also noted systemic barriers to including WG availability in school meals. While both schools are responsible for designing their lunch menus, they typically outsource food preparation and ingredient sourcing to external contractors. As a result, introducing more WG options requires coordination and agreement with these contractors. Findings from a Minnesota school illustrate similar challenges, in which successful implementation depended on contractors fully understanding the school’s requirements and the school clearly articulating its plans for WG inclusion [32]. These findings are consistent with a recent scoping review, which identified that school food procurement contracts and contractor priorities can influence the availability, quality, and consistency of healthier menu items, including WG [33]. Participants noted that negotiations are further complicated by budget constraints, as the higher cost of WG relative to refined grains limits flexibility within government-provided school lunch budgets. Studies suggest that private-sector contractors may prioritize profitability over healthier options [32–34]. This may limit the supply, quality, or use of WG in meals. Additionally, participants observed that Thai School Lunch, a platform used by schools to design and report lunch menus to district authorities, is highly restrictive and inflexible [35]. The system offers a limited number of menu templates, with only a narrow set of customization options. This lack of flexibility poses a unique challenge in the Thai elementary school context, which has not been widely documented in prior literature on WG promotion in schools. It is worth noting that many Thai elementary schools prepare meals in-house, which may involve system-level challenges not captured in this study [36]. Identifying strategies to navigate or improve such system-level barriers would therefore represent an important area for future research and policy development.

A further barrier to increasing WG provision is the lack of incentives at the school level. Participants reported that many schools are not fully aware of the nutritional benefits of WG, as reflected in teachers’ limited knowledge of their health impacts. Additionally, WG is absence from Thai “nutritional flag” and the Thai FBDGs, the official nutrition reference used for school lunch planning, simply encourage the consumption of brown rice as the main energy source with no definite quantity [10,37], unlike the American or Malaysian dietary guidelines that specifically require schools to serve WG at least half of the grain portion [38]. Therefore, there is currently no regulation or policy mandating the inclusion of WG in elementary school lunches, nor are there incentives to encourage schools to do so. With leniency in regulations, many studies in Thailand have found that schools continue to use white rice as the primary staple, and to date, none have reported substantial use of brown rice in schools [39,40]. This lack of top-down guidance and structural support contributes to the low prioritization of WG in school meal planning. To provide schools with both a clear benchmark and justification for increasing WG offerings, quantifying the specific portion of WG recommended to Thai FBDGs was proposed as a practical step to institutionalize WG promotion.

Additionally, participants suggested that the Ministry of Public Health take a more active role in raising awareness about WG and their health benefits. Through a public–private partnership integrating research, PR, and national dietary guidelines, the Danish Whole Grain Partnership successfully increased WG consumption in Denmark by approximately 75% over a 15-year period [41,42]. A participant emphasized that awareness efforts should particularly target key program influencers within schools, including teachers responsible for delivering nutrition education, those involved in menu design, and members of the school’s administrative leadership.

Barriers to whole grain procurement in Thai elementary schools extend well beyond nutritional concerns, the focus of our study. Thailand is one of the world’s largest rice exporters, and shifts in domestic procurement policies carry significant implications for trade income and for farmers operating on precarious profit margins. Transitioning to alternative varieties, such as brown rice, presents additional challenges, including a shorter shelf life and limited integration into national agricultural infrastructure, which partly explains why white rice has remained the dominant staple despite decades of investment in rice science and technology. Future research should therefore engage economists, agricultural specialists, and development policy experts to provide a more holistic feasibility assessment, as any meaningful multisectoral response would require flexibility, long-term commitment, and coordination across schools, farmers, distributors, and policymakers [43].

### Proposed Strategies

To address student resistance, participants proposed several strategies, including school-based education on WG and their benefits, as well as hands-on activities such as cooking classes and taste-testing sessions. Participants perceived elementary-aged children as relatively open to trying new foods in supportive settings, suggesting a window of opportunity to introduce WG during this developmental stage. We conducted a concurrent survey with the 4-6^th^ graders at the elementary schools included in this interview study and found that children were in fact willing to try and eat more whole grains, especially if they have health benefits (unpublished work). This perception is supported by studies in various settings indicating that familiarity with new foods can be increased through structured, low-pressure exposure. Sensory-based food education approaches, such as the French Classes du Goût program, demonstrate that child-centered activities engaging multisensory experiences can reduce food neophobia and promote willingness to try unfamiliar foods [44–46]. In addition, peer influence and teacher modeling have been shown to facilitate the acceptance of new foods among children aged 8–12 years, further reinforcing openness to unfamiliar foods when introduced in socially supportive contexts [44].

Participants suggested that schools explore alternative funding sources, such as contributions from parent organizations or local community partners, to help offset the cost of WG products. Similar approaches have been used in Malaysia, where parent-funded school meal programs, such as Hidangan Berkhasiat di Sekolah, support the provision of nutritionally guided breakfasts. However, such funding is often uneven and time-limited and may contribute to disparities in food provision across schools [47]. For sustainable, system-wide change, participants emphasized the importance of government-level financial support, including increased per-meal allocations or targeted subsidies for the procurement of WG. International experience with government-funded school meal programs suggests that such investment can improve both meal quality and child nutrition [47–49]. Future efforts should consider how temporary external funding can be complemented by government-level financing and policy mechanisms to support sustained WG provision in Thai elementary schools. Additionally, WG options in Thai school lunches, if any, are currently largely limited to brown rice and whole wheat bread. Participants suggested that exploring a broader range of affordable WG could help schools design more feasible and cost-effective menus. For example, locally available and affordable WG such as pigmented rice (Riceberry, Hom Nil, Sangyod, Red Hom Mali, and black glutinous rice), whole wheat, oat, corn, job’s tear, could be introduced to diversify menu options and lower costs [8]. Achieving this, however, requires collaboration beyond the nutrition sector, particularly with economic and agricultural stakeholders, given Thailand’s deeply embedded investment in rice production.

### Strengths and Limitations

This study included both key stakeholders and experts in the nutrition field, enabling a more comprehensive understanding of the barriers and facilitators to WG provision in Thai elementary schools. A rigorous, iterative coding process was employed, with two independent coders to minimize bias. The matrixing process among coders yielded high agreement, and most themes were applied consistently, demonstrating the codebook’s strength and reliability. While the study provided a thorough understanding of students’ attitudes toward WG, insights into students’ barriers were based solely on key adults’ perceptions and may not fully reflect students’ actual views. Additionally, the study was conducted in only two elementary schools within a single province of Thailand. Both schools had an existing partnership with the university to promote student health through their lunch programs and were located in central Thailand, a region where white rice is predominantly consumed. As a result, these schools may not be representative of the broader population of Thai elementary schools, particularly those with different structures or without similar partnerships, which may limit the transferability of the findings.

### Implications

Future research should include a larger and more diverse sample of Thai elementary schools, including those using different meal provision models and those in different regions who may have different rice consumption patterns and varieties, to improve the generalizability of findings across the school system. Importantly, the present findings are based on adult perceptions of children’s attitudes. Direct engagement with students through interviews or focus groups would provide critical insight into children’s preferences and inform more targeted intervention strategies. In addition, future research should identify existing strengths within Thai elementary schools that already support WG inclusion and examine how these assets can be leveraged to facilitate implementation. Finally, given the roles of both households and government identified in this study, further work should explore opportunities for coordinated, multi-sector collaboration to support sustained increases in WG intake among Thai elementary school children.

## Conclusion

WG consumption remains low among Thai schoolchildren, despite rising rates of childhood obesity and strong global emphasis on WG intake as part of healthy dietary patterns. To the best of our knowledge, this is the first study to examine the limitations and facilitators of WG provision in Thai elementary schools from the perspective of adult stakeholders. Multiple barriers were identified, including perceived student resistance stemming from limited exposure and knowledge, unique structural constraints within school meal systems, and the absence of clear policy mandates. Despite significant reports of student resistance toward WG, these findings were based only on adults’ perceptions. Assessing schoolchildren’s knowledge, attitudes, and behaviors regarding WG is needed to validate these findings. Future research should include schools from a wider range of settings, including schools with different meal preparation models and those with existing WG provision. Such diversity would support a more comprehensive understanding of context-specific challenges within Thai elementary schools.

Success in increasing WG consumption among children requires both behavioral change and a supportive resource environment. Therefore, collaboration among multisectoral stakeholders, including schools, households, food service providers, and government agencies, is essential. Schools, along with food service providers, are encouraged to explore the local WG options that align with current budgets and students’ preferences. In parallel, policymakers need to be informed of the importance of WG intake among children and initiate top-down support for WG procurement in schools.

## Data Availability

Due to the sensitive and potentially identifying nature of the qualitative interview data, the full de-identified transcripts cannot be made publicly available. De-identified data may be made available to qualified researchers upon reasonable request. Illustrative excerpts supporting the reported themes are provided within the manuscript.

## Acknowledgements

The authors wish to thank the peers, colleagues, students, and teachers involved in this project for their valuable input and ongoing support. We are especially grateful to the participating elementary schools, particularly the teachers, staff, and chefs, as well as the INMU faculty and staff, who provided important insights and perspectives. We also thank Joel Gittleson for his mentorship on manuscript development, and the writing of seminar peers for their thoughtful feedback and critical discussions.

## Supporting information

**S1 Table. Codebook detailing the initial 7 themes and subthemes, with definitions used for the Framework analysis, prior to consolidation into the final 5 themes reported in the manuscript.**

**S2 Table. Completed SRQR checklist for transparent reporting of this qualitative study.**

**S1 File. Human participants research checklists.**

**S2 File. Certificate of approval, Mahidol University Central Institutional Review Board.**

**S3 File. IRB Approval Memo, Johns Hopkins Bloomberg School of Public Health Institutional Review Board Office.**

